# Sevoflurane-based enhancement of phase-amplitude coupling and localization of the epileptogenic zone

**DOI:** 10.1101/2021.10.21.21265330

**Authors:** Keiko Wada, Masaki Sonoda, Ethan Firestone, Kazuki Sakakura, Naoto Kuroda, Yutaro Takayama, Keiya Iijima, Masaki Iwasaki, Takahiro Mihara, Takahisa Goto, Eishi Asano, Tomoyuki Miyazaki.

## Abstract

**Objective:** Phase-amplitude coupling between high-frequency (≥150 Hz) and delta (3-4 Hz) oscillations - modulation index (MI) - is a promising, objective biomarker of epileptogenicity. We determined whether sevoflurane anesthesia preferentially enhances this metric within the epileptogenic zone.

**Methods:** This is an observational study of intraoperative electrocorticography data from 621 electrodes chronically implanted into eight patients with drug-resistant, focal epilepsy. All patients were anesthetized with sevoflurane during resective surgery, which subsequently resulted in seizure control. We classified ‘removed’ and ‘retained’ brain sites as epileptogenic and non-epileptogenic, respectively. Mixed model analysis determined which anesthetic stage optimized MI-based classification of epileptogenic sites.

**Results:** MI increased as a function of anesthetic stage, ranging from baseline (i.e., oxygen alone) to 2 minimum alveolar concentration (MAC) of sevoflurane, preferentially at sites showing higher initial MI values. This phenomenon was accentuated just prior to sevoflurane reaching 2 MAC, at which time, the odds of a site being classified as epileptogenic were enhanced by 86.6 times for every increase of 1 MI.

**Conclusions:** Intraoperative MI best localized the epileptogenic zone immediately before sevoflurane reaching 2 MAC in this small cohort of patients.

**Significance:** Prospective, large cohort studies are warranted to determine whether sevoflurane anesthesia can reduce the need for extraoperative, invasive evaluation.

**Highlights:**

- We measured the modulation index on intraoperative electrocorticography recording.
- Sevoflurane enhanced the modulation index differentially across the epileptogenic and non- epileptogenic sites.
- The modulation index best discriminated these two groups of sites before sevoflurane reached 2 minimum alveolar concentration.

## 1. Introduction

Interictal spike-and-wave discharges are valuable biomarkers to estimate the epileptogenic zone and guide surgical resection for patients with drug-resistant focal epilepsy (Palmini et al., 1995; Rosenow and Lüders, 2001). Each spike is generally accompanied by broadband, high-frequency activity at ≥150 Hz and a 3-4 Hz slow wave (Fürbass et al., 2020; Jacobs et al., 2011). As a surrogate marker reflecting the severity of these epileptiform discharges, modulation index (MI) quantifies the coupling between >150 Hz high-frequency amplitude and the phase of 3-4 Hz slow waves, and investigators suggest it can localize the seizure onset zone (Iimura et al., 2018; Nariai et al., 2020; Nonoda et al., 2016). Furthermore, resection of sites showing increased MI, measured on extraoperative electrocorticography (ECoG) during slow-wave sleep, predicted successful seizure control (Kuroda et al., 2021; Motoi et al., 2018). The logical next step is to determine the utility of *<u>intraoperatively measured</u>* MI in epilepsy presurgical evaluation.

To that point, the present study aimed to determine whether an acute, sevoflurane-induced increase in MI during intraoperative ECoG recording will localize epileptogenic sites and identify the optimal anesthetic stage for this purpose. Since all patients achieved International League Against Epilepsy (ILAE) class 1 outcomes, we defined the epileptogenic sites as those included in the resection because postoperative seizure control infers that the entire epileptogenic zone was removed (Wieser et al., 2001); previously, investigators have suggested that only resecting the seizure onset zone may fail to control seizures in a subset of patients, as other parts of the epileptogenic zone remain intact (Jehi, 2018; Rosenow and Lüders, 2001; Singh et al., 2015).

In the present study, all patients had extraoperative ECoG recording with subdural electrodes in an epilepsy monitoring unit, to capture and characterize habitual seizures; they then had intraoperative ECoG recording during a second surgery for electrode explantation and cortical resection. As part of standard care anesthetization during the resection, we had a rare opportunity to measure the effects of dynamically *<u>rising</u>* sevoflurane concentration – zero to 2 minimum alveolar concentration (MAC) - on ECoG signals at epileptogenic and non-epileptogenic sites (García-Tejedor et al., 2020; Iijima et al., 2000). Investigators previously reported that sevoflurane increased the occurrence rate of interictal spike discharges (Kurita et al., 2005; Shimizu, 1997; Tanaka et al., 2017; Watts et al., 1999) and high-frequency events (Orihara et al., 2020), in a dose-dependent manner. Alternatively, some suggest that too high concentration of sevoflurane may induce sharp transients and high-frequency discharges unrelated to the epileptogenicity that may mimic spike-and-wave discharges (Hisada et al., 2001; Orihara et al., 2020; Tanaka et al., 2017). In these sevoflurane-related studies (Kurita et al., 2005; Orihara et al., 2020; Tanaka et al., 2017; Watts et al., 1999), *<u>discrete</u>* spikes and high-frequency activity were detected based on visual counting, which is time-consuming and suffers from suboptimal inter-rater agreement. In contrast, the present study used an open-source toolbox (Miyakoshi et al., 2013) and objectively quantified MI as a *<u>continuous</u>* measure (Kuroda et al., 2021). Since MI is fundamentally derived from spike-and-wave signals and previous reports demonstrate sevoflurane’s ability to augment spike and high-frequency occurrence rates, we expected that MI would increase as a function of the sevoflurane anesthetic stage. The specific question to be addressed was whether the magnitude of sevoflurane-related MI augmentation was higher at cortical sites showing increased MI during the non-anesthetized baseline period. We then determined what anesthetic stage optimized the accuracy of MI-based classification of epileptogenic and non-epileptogenic sites.

## 2. Methods

### 2.1. Patients

This retrospective, observational study was approved by the Institutional Ethical Committee at the National Center of Neurology and Psychiatry (NCNP), Tokyo, Japan (approval number: A2021-050). This is a secondary analysis using data from the project titled, “Exploratory clinical study to identify epileptogenic foci by statistical analysis of electrocorticographic signals during general anesthesia in patients with intractable epilepsy (observational study)”. We obtained written consent from all patients or legal guardians, for those under the age of 18 or incapable of providing written approval.

We studied eight patients with drug-resistant, focal epilepsy (age range: 4-22 years) who underwent resective surgery from October 2018 to March 2020 and met the eligibility criteria. The inclusion criteria were as follows: [1] the patients underwent extraoperative ECoG and subsequent focal resection at the NCNP, [2] the patients met American Society of Anesthesiologists Physical Status 1 or 2, reflecting good perioperative health status; (Aronson et al., 2003), and [3] the patients achieved International League Against Epilepsy (ILAE) class 1 outcomes at least 12 months after surgical resection (Wieser et al., 2001). The exclusion criteria consisted of the following: [1] patients had contraindication to sevoflurane or propofol, and [2] patients took anesthetic agents other than sevoflurane, propofol, remifentanil, or rocuronium during the intraoperative ECoG analysis period described in more detail below. **Table 1** shows the full patient profile.

**Table 1:** Patient profile.

| Patient number | Sex | Antiepileptic drugs | Number of electrodes | Resection area | Pathology |
| --- | --- | --- | --- | --- | --- |
| 1 | F | TPM, VPA | 75 | Left frontal | Gliosis |
| 2 | M | CBZ, LEV, LTG, PER | 63 | Left occipital | Ulegyria |
| 3 | F | LEV, LTG, VPA, | 120 | Right frontal and parietal | Polymicrogyria |
| 4 | F | CBZ, CLB, TPM | 114 | Right temporal and parietal | Gliosis |
| 5 | M | ZNS, LEV, CLB | 41 | Left frontal and temporal | FCD type IIb |
| 6 | M | CBZ, VPA, LTG | 62 | Right frontal | FCD type IIa |
| 7 | M | CLB, LEV, LCM, PB | 62 | Right frontal | FCD type IIb |
| 8 | M | CBZ, LCM | 84 | Right temporal | Gliosis |

AEDs = Antiepileptic drugs; CBZ = Carbamazepine; CLB = Clobazam; F = Female; FCD = Focal cortical dysplasia; iEEG = intracranial electroencephalography; IFG = Inferior frontal gyrus; LCM = Lacosamide; LEV = Levetiracetam; LTG = Lamotrigine; M = Male; PB = Phenobarbital; PER = Perampanel; TPM = Topiramate; VPA = Valproic acid; ZNS = Zonisamide.

### 2.2. Intracranial electrode placement

For all patients, we placed platinum disk electrodes (10 mm center-to-center distance) in combination with depth electrodes (5 mm center-to-center distance) on the affected hemisphere to localize the boundary between the seizure onset zone and eloquent areas; it should be noted that patient #2 only received disk electrodes (**Table 1**). The spatial extent of intracranial electrode placement was determined strictly based on clinical needs and guided by an interdisciplinary team of physicians integrating collective evidence from semiology, scalp video-EEG, anatomical, and functional neuroimaging findings (Takayama et al., 2021, 2020).

### 2.3. Extraoperative electrocorticography (ECoG) recording and electrical stimulation mapping

At the inpatient ward, we performed continuous video-ECoG recording with a sampling rate of 1,000 Hz and a band-pass filter of 0.016 to 300 Hz, for invasive evaluation of the epileptogenic brain regions. For each patient, we generated a three-dimensional magnetic resonance imaging (MRI) surface image with intracranial electrodes superimposed on their anatomically accurate locations, as previously reported (Ikegaya et al., 2019; Nakai et al., 2017). Oral anti-seizure medications were tapered in patients #1, #3, #4, and #8 (**Table 1**) to increase the chance of capturing seizure events as per the treating physician’s discretion. In patients #3, #4, and #8, all anti-seizure medications were resumed at least two days before the resective surgery. In patient #1, only Valproic acid was discontinued, and it was not resumed before the resective surgery. At least two days before the resective surgery, all but patient #1 underwent electrical stimulation mapping to localize the eloquent areas (Takayama et al., 2021, 2020). Habitual seizures were captured in all eight patients, and a board-certified epileptologist (M.I.) localized the seizure onset zone (Asano et al., 2009).

### 2.4. Intraoperative ECoG under general anesthesia during the resection surgery

After extraoperative ECoG recording to capture and characterize habitual seizures, each patient was transferred to the operating room for their focal, resective surgery. All procedures were part of standard clinical practice and approved by the Institutional Ethics Commission at the NCNP. Each patient took anti-seizure medications but no other premedication on the morning of the resective surgery (**Table 1**). Before general anesthesthetic induction, we initiated ECoG recording and employed standard monitoring, including percutaneous oximeter saturation, non-invasive blood pressure measurement, and electrocardiogram. The same electrode arrays were used for both extraoperative and intraoperative ECoG recordings. General anesthesia was then rapidly induced using propofol (2.16 mg/kg on average across patients), immediately followed by analgesia with remifentanil (2.06 mcg/kg on average) and muscle relaxation with rocuronium. Next, sevoflurane was initiated, patients were intubated with an endotracheal tube, and sevoflurane concentration was dynamically increased from zero to 2.0 MAC; we continued ECoG recording for an additional three minutes, at this maintenance level. Throughout the intraoperative ECoG recording period, each patient was also continuously given 100% oxygen. The aforementioned procedure, including 2.0 MAC sevoflurane, is suggested to be within a clinically acceptable range (Malan et al., 1995; Iijima et al., 2000; Kurita et al., 2005). **Table 2** shows the dose of anesthetic agents given to each patient.

**Table 2:** Anesthesia information.

|  |  |
| --- | --- |
| Number of patients | 8 |
| Age median | 11.5 years |
| (range, IQR) | (4–22, 9.5–15.3) |
| BMI median | 17.8 kg/m<sup>2</sup> |
| (range, IQR) | (16.6–30.9, 16.7–19.1) |
| Propofol dose at induction of anesthesia median | 2.07 mg/kg |
| (range, IQR) | (2.00–2.67, 2.00–2.21) |
| Remifentanyl dose at induction of anesthesia median | 2.00 mcg/kg |
| (range, IQR) | (1.95–2.29, 2.00–2.07) |
| P<sub>ET</sub>CO<sub>2</sub> on average* median | 35 |
| (range, IQR) | (26–38, 34–37) |
| Blood pressure on average† median | 70 mmHg |
| (range, IQR) | (61–82, 67–77) |
| Heart rate on average† median | 82 beats per minute |
| (range, IQR) | (70–102, 77–88) |
| Body temperature on average* median | 36.5 °C |
| (range, IQR) | (35.7–36.7, 36.3–36.7) |
| Duration to reach sevoflurane 2.0 MAC | 6.8 minutes |
| (range, IQR) | (4.8–11.4, 5.3–7.5) |

\*, Values averaged over the period when sevoflurane was maintained at 2.0 MAC.
†, Values averaged over the period of intraoperative electrocorticography recording.
BMI = Body mass index; °C = Degrees Celsius; IQR = Interquartile range; MAC = Minimum alveolar concentration; P<sub>ET</sub>CO<sub>2</sub> = Partial pressure of exhaled carbon dioxide.

### 2.5. Defining the five anesthetic stages

The intraoperative ECoG epoch was divided into the following five anesthetic stages: [1]’ *baseline wakefulness’* before anesthetic induction; next, the time period between sevoflurane initiation and reaching 2.0 MAC divided into [2] *‘first 1/3’*; [3] *‘middle 1/3’*; [4] *‘last 1/3’*; and [5] *‘sevoflurane 2*.*0 MAC maintenance’*. On average, the duration of ‘*baseline wakefulness*’, ‘*first 1/3*’, ‘*middle 1/3*’, ‘*last 1/3*’, and ‘*2*.*0 MAC maintenance*’ stages were 4, 2, 2, 2, and 4 minutes, respectively. We assumed that the sevoflurane concentration increased as a function of the anesthetic stage. We determined during which anesthetic stage the MI best discriminated between the epileptogenic and non-epileptogenic sites below.

### 2.6. Resective surgery

Resection areas included the epileptologist-defined seizure onset zone and neighboring, MRI-visible cortical lesions; whereas the eloquent areas were maximally preserved (Takayama et al., 2021, 2020). Pertinently, the MI results mentioned below were not involved in deciding the location and bounds of resection.

### 2.7. Definition of the epileptogenic sites for retrospective analysis

The present study defined the epileptogenic areas as electrode sites included in the resection, confirmed by intraoperative photographs. We have previously reported that the spatial extent of focal resection defined by intraoperative photographs was highly concordant with that estimated by postoperative MRI (Kuroda et al., 2021).

### 2.8. Computation of modulation index (MI)

We only included artifact-free ECoG epochs and channels in the subsequent analysis. Initially, investigators (E.F. and N.K.) blinded to the epileptogenic classification of all electrodes exported ECoG signals on a bipolar montage; these signals were then fed into the open-source EEGLAB Toolbox winPACT (https://sccn.ucsd.edu/wiki/WinPACT) to quantify MI for all sites and anesthetic stages (Miyakoshi et al., 2013). Thereby, MI reflects the strength of the association between the amplitude of high-frequency activity at ≥150 Hz and the instantaneous phase of a local slow wave at 3-4 Hz, averaged across all ECoG time points within each anesthetic stage (Kuroda et al., 2021; Motoi et al., 2018).

### 2.9. Statistical analysis

[Aim 1] Testing our hypothesis that sevoflurane increases MI values in a stage-dependent manner, we used mixed model analysis to determine the relationship between the sevoflurane anesthetic stage and MI, at all electrode sites. We treated the sevoflurane anesthetic stage as a fixed-effect, ordinal, predictor variable, patient and intercept as random-effect factors, and MI, at a given electrode site during a given anesthetic stage, as the outcome measure. A fixed-effect estimate greater than zero suggests that more advanced sevoflurane stages were associated with increased MI. We considered a p-value of <0.05 as significant and performed the computation using MATLAB Statistical Toolbox (Mathworks, Natick, MA, USA).

Furthermore, we determined whether a given sevoflurane anesthetic stage increased MI preferentially at electrode sites with higher baseline MI values. To address this question, mixed model analysis incorporated the interaction between the baseline MI value and sevoflurane anesthetic stage as a fixed-effect predictor. The random-effect and outcome variables were identical to those above, and similarly, a fixed-effect estimate greater than zero suggests that a more advanced sevoflurane stage increased MI preferentially at sites with higher baseline MI values.

[Aim 2] The logistic mixed model analysis determined what anesthetic stage optimized the accuracy of MI-based classification of epileptogenic and non-epileptogenic sites. For each anesthetic stage, we treated a given MI value at a given electrode site as a fixed-effect predictor, patient and intercept as random-effect factors, and epileptogenicity as the outcome measure. We repeated this logistic mixed model analysis for the five anesthetic stages and considered a p-value of <0.01 as significant.

Finally to assess the accuracy of the MI-based logistic mixed model in localizing epileptogenic sites, we calculated the receiver operating characteristics (ROC) curve (Asano et al., 2009; Kuroda et al., 2021), using the entire pool of 621 electrode sites from all eight patients.

## 3. Results

### 3.1. Anesthetic management during intraoperative ECoG recording

In general, no patients experienced adverse events during the intraoperative ECoG recording period nor needed cardioactive drugs. Blood pressure, heart rate, body temperature, and oxygen saturation levels remained within normal limits. The only anomalies: the youngest patient (Patient #7) received prophylactic atropine to prevent excessive bradycardia, and patient #4 was maintained hypocapnic, whereas the remaining patients were normocapnic. On average, it took 6.9 minutes for the end-tidal sevoflurane concentration to reach 2.0 MAC.

### 3.2. [Aim 1] Effect of the sevoflurane anesthetic stage on modulation index (MI)

The mixed model analysis demonstrated that more advanced sevoflurane anesthetic stages were associated with increased MI (fixed effect estimate: +0.013; 95% confidence interval [CI]: +0.012 to +0.015; p-value: <0.001; t-value: 15.82; degree of freedom [DF]: 3040; **Figure 1**). This observation indicates that for each increase of the anesthetic stage, the overall, raw MI value increased by 0.013, across all eight patients.

**Figure 1.**
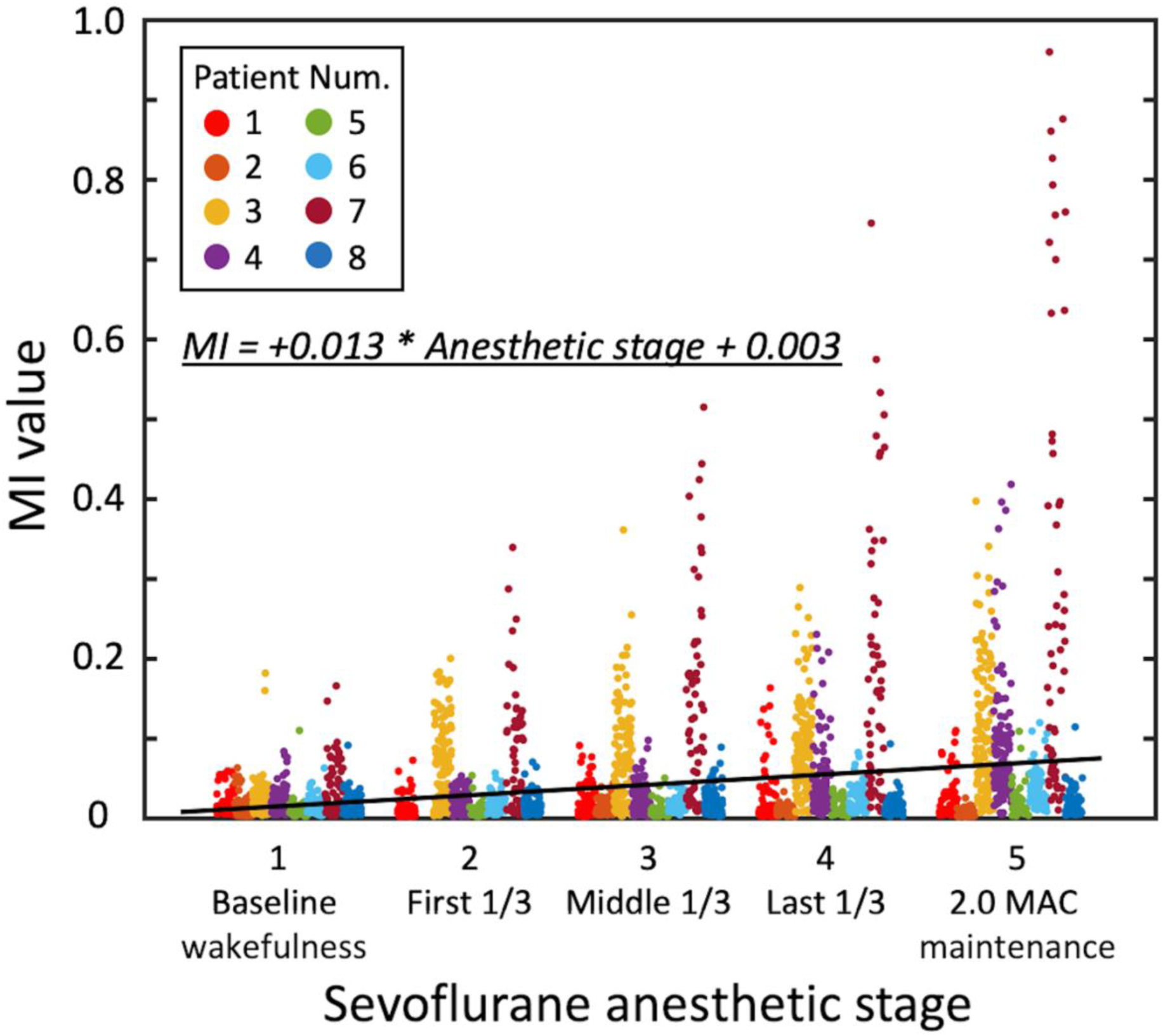
Effect of the sevoflurane anesthetic stage on modulation index (MI). The scatter plot shows MI values at given electrode sites during each sevoflurane anesthetic stage. Individual patients are color-coded. The solid line reflects the regression line estimated by the mixed model analysis. The sevoflurane anesthetic stage was incorporated into the mixed model as an ordinal variable. Num. = number. MAC = minimum alveolar concentration.

Furthermore, mixed model analysis incorporating the interaction between anesthetic stage and baseline MI values suggested that more advanced sevoflurane stages increased MI preferentially at electrode sites with higher baseline MI values (fixed effect estimate of the interaction: +0.55; 95% CI: +0.48 to +0.62; p-value: <0.001; t-value: 14.90; DF: 3038; **Table 3**).

**Table 3:** Mixed model analysis to determine the relationship between the anesthetic stage and modulation index (MI).

| Variables | Estimate | 95% CI |  | p |
| --- | --- | --- | --- | --- |
|  |  | LL | UL |  |
| Interaction b/w baseline MI value and sevoflurane anesthetic stage | 0.55 | 0.48 | 0.62 | <0.001 |
| Baseline MI value | -0.28 | -0.52 | -0.032 | 0.027 |
| Sevoflurane anesthetic stage | 0.0014 | -0.00077 | 0.0035 | 0.21 |

The sevoflurane anesthetic stage was treated as an fixed effect ordinal predictor variable.
Increased interaction between (b/w) the baseline MI value and sevoflurane anesthetic stage was associated with increased MI at a given electrode site. In other words, more advanced sevoflurane anesthetic stages augmented MI preferentially at sites with higher baseline MI values. The degrees of freedom = 3038.
CI = Confidence interval; LL = Lower limit; MI = Modulation index; UL = Upper limit.

### 3.3. [Aim 2] Optimal anesthetic stage for classification of epileptogenic sites

The logistic mixed model analysis indicated that the *‘last 1/3’* anesthetic stage optimized the MI-based classification of epileptogenic and non-epileptogenic sites. For each increase of 1 MI during the *‘last 1/3’* anesthetic stage, the odds of a given site being classified as epileptogenic increased by 86.6 times; in other words, for each increase of 0.2 MI, the odds increased by 2.44 times. The fixed effect estimates (odds ratio; p-value) of the local MI values were -2.04 (0.13; 0.44) during the *‘first 1/3’*, +2.90 (18.2; 0.10) during the *‘middle 1/3’*, +4.46 (86.6; 0.003) during the *‘last 1/3’*, and +1.38 (3.97; 0.11) during the *‘2*.*0 MAC maintenance’* stage (**Table 4**; **Figure 2**).

**Table 4:** Logistic mixed model-based classification of the epileptogenic and non-epileptogenic sites.

| Variables | Odds | 95% CI |  | p |
| --- | --- | --- | --- | --- |
|  |  | LL | UL |  |
| <u>'Baseline wakefulness':</u> $r^2 = 0.10$, $df = 619$ | | | | |
| MI value | 11.91 | 0.001 | 1.1×10<sup>5</sup> | 0.59 |
| <u>'First 1/3':</u> $r^2 = 0.12$, $df = 556$ | | | | |
| MI value | 0.13 | 7.25×10<sup>-4</sup> | 23.34 | 0.44 |
| <u>'Middle 1/3':</u> $r^2 = 0.11$, $df = 619$ | | | | |
| MI value | 18.21 | 0.58 | 8.10×10<sup>7</sup> | 0.10 |
| <u>'Last 1/3':</u> $r^2 = 0.14$, $df = 619$ | | | | |
| **MI value** | **86.58** | **4.48** | **1.67×10<sup>3</sup>** | **0.003** |
| <u>'2.0 MAC maintenance':</u> $r^2 = 0.11$, $df = 619$ | | | | |
| MI value | 3.97 | 0.73 | 21.54 | 0.11 |

For each increase of 1 MI during the 'last 1/3' anesthetic stage, the odds of a given site being classified as epileptogenic increased by 86.6. Significant predictor ( $p < 0.01$ ) is denoted in **bold** typeface.
CI = Confidence interval; df = degree of freedom; LL = Lower limit; MAC = Minimum alveolar concentration; MI = Modulation index; $r^2$ = coefficient of determination; UL = Upper limit.

**Figure 2.**
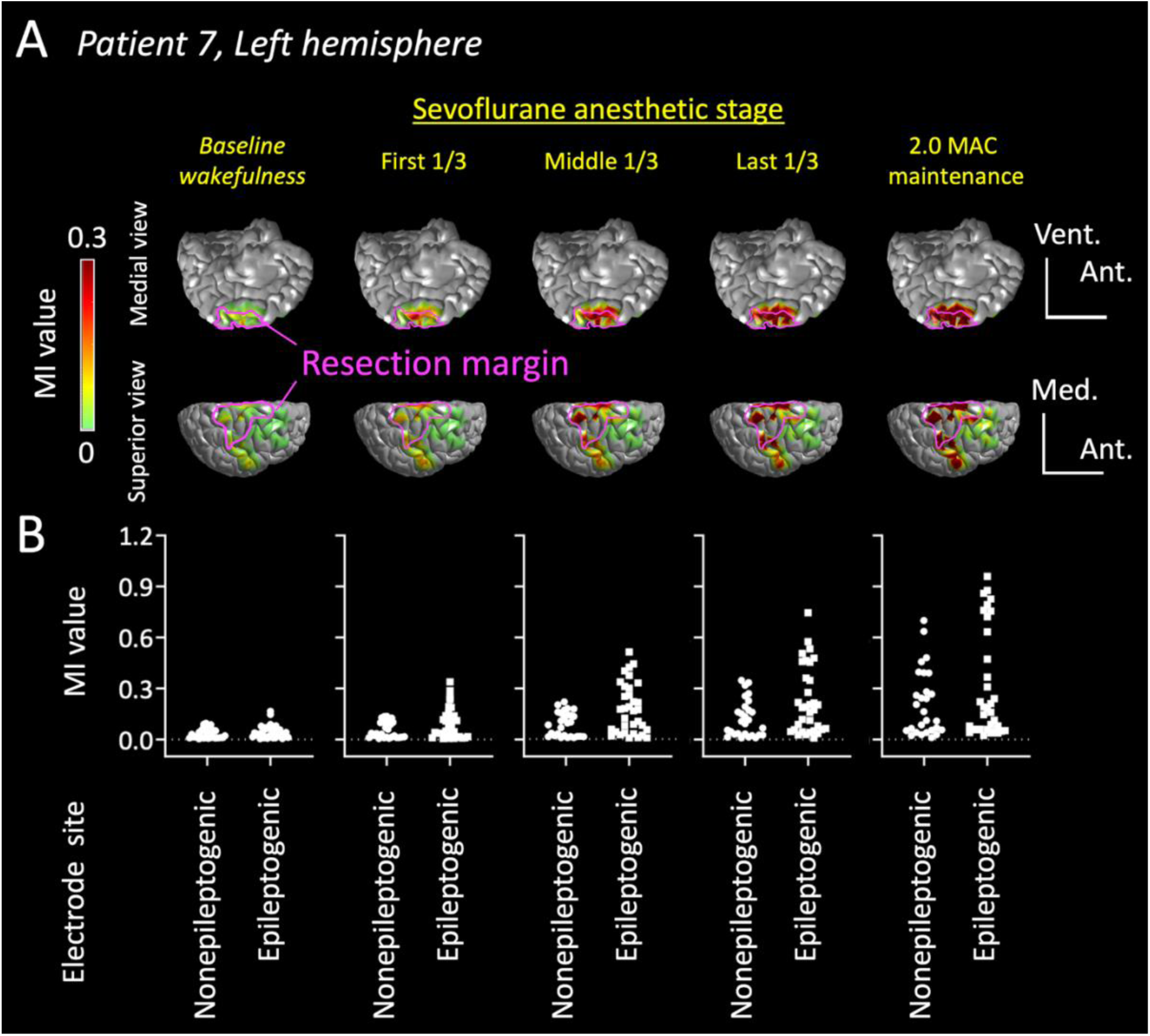
Spatial relationship between the epileptogenic sites and modulation index (MI) during sevoflurane anesthetic stages. (A) The three-dimensional brain surface images present the relationship between the resection margin and MI in each anesthetic stage in patient #7. MI at the epileptogenic sites was increased minimally during the ‘*baseline wakefulness*’ but prominently during the ‘*last 1/3*’ stage. During the ‘*2*.*0 MAC maintenance*’ period, MI was increased not only at the epileptogenic sites but also at some of the non-epileptogenic sites. Pink lines denote the resection margin. Ant. = anterior; Med. = medial; Vent. = ventral. (B) The scatter plots denote MI values at the epileptogenic and non-epileptogenic sites.

Assessing model accuracy, the ROC analysis demonstrated that MI during the *‘last 1/3’* anesthetic stage localized the epileptogenic site with an area under the curve of 0.71 (bootstrap 95% confidence interval: +0.65 to +0.77, n=5,000). Thereby, electrode sites with a MI-based model probability above 0.45 localized the epileptogenic sites with a specificity of 0.95 and a sensitivity of 0.23 (**Figure 3**).

**Figure 3.**
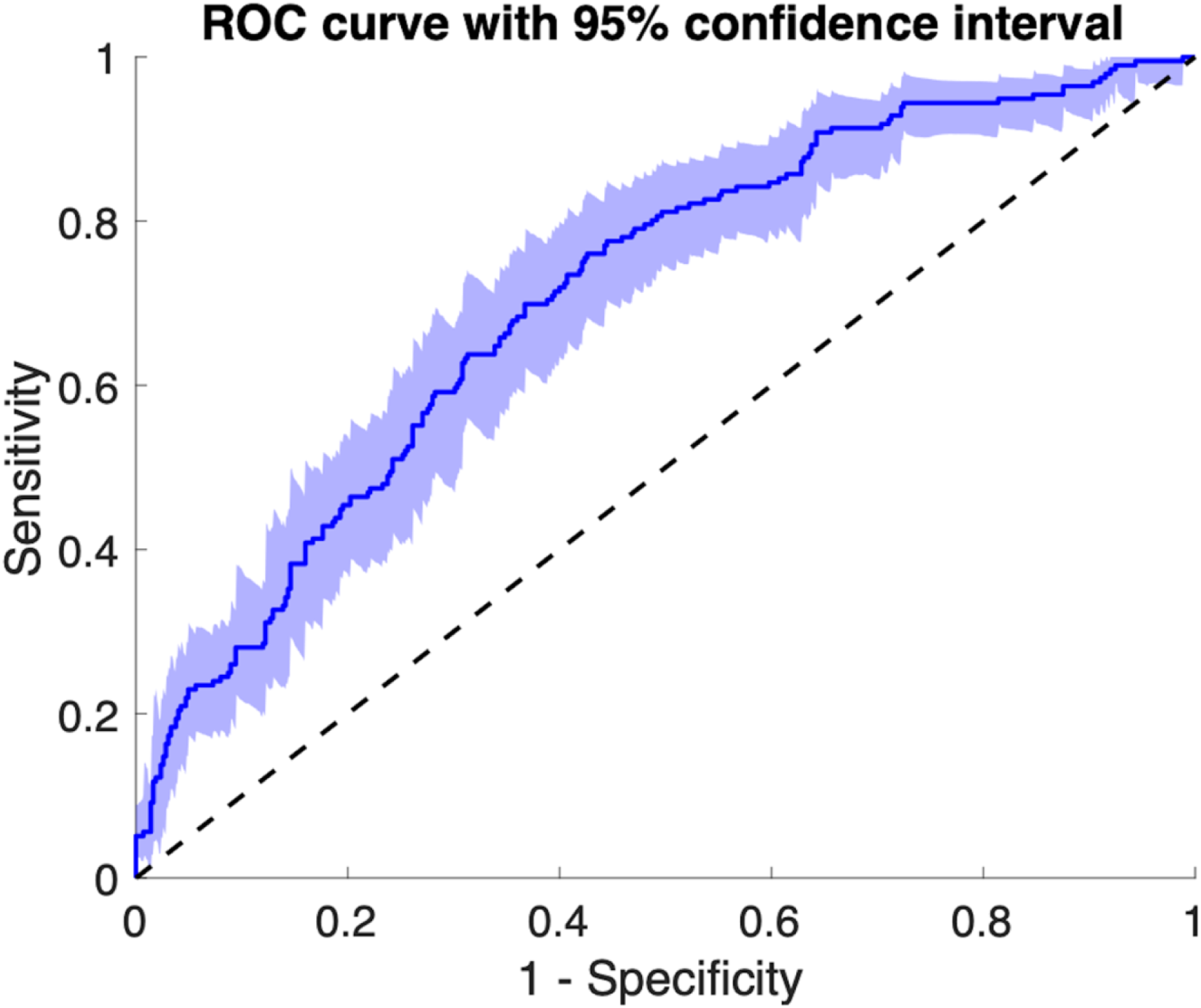
Receiver operating characteristics (ROC) analysis. The ROC curve indicates the accuracy of the modulation index (MI)-based logistic mixed model in localizing the epileptogenic sites during the *‘last 1/3’* anesthetic stage. The area under the ROC curve was 0.71 (bootstrap 95% confidence interval: +0.65 to +0.77). The number of bootstrap repetitions was set to 5,000.

## 4. Discussion

### 4.1. Significance

To our knowledge, this is the first study to demonstrate the potential utility of intraoperatively measured phase-amplitude coupling for localization of the epileptogenic zone, in patients undergoing epilepsy surgery. The MI - quantifying phase-amplitude coupling between high-frequency activity at ≥150 Hz and 3-4 Hz slow-waves - increased preferentially at the epileptogenic sites, as a function of sevoflurane anesthetic stage. This phenomenon was optimized during the two-minute period prior to sevoflurane reaching 2.0 MAC (i.e., *‘last 1/3’* stage).

Our results are consistent with those reported in previous studies of extraoperative and intraoperative ECoG recordings. For example, an extraoperative ECoG study of 135 patients reported that resection of sites showing increased MI during slow-wave sleep predicted successful seizure control after surgery (Kuroda et al., 2021). An intraoperative ECoG study of 12 patients reported that visually-identified spike discharges were more frequent during 1.5 MAC sevoflurane, compared to the same concentration of isoflurane anesthesia (Watts et al., 1999). Likewise, another intraoperative ECoG study of 13 patients demonstrated that interictal spike discharges were more frequent and extensive during 1.5 MAC sevoflurane than 0.5 MAC sevoflurane anesthesia (Kurita et al., 2005), and they were able to localize the seizure onset zone with a sensitivity of 0.91 and specificity of 0.81 (Kurita et al., 2005). Looking at high-frequency activity, a recent intraoperative ECoG study of seven patients with mesial temporal lobe epilepsy visually counted high-frequency events at medial and lateral temporal lobe sites during sevoflurane anesthesia (Orihara et al., 2020). Raising sevoflurane from 1.5% to 3.0% in half percent increments (presumably equivalent to ‘from 0.8 to 1.7 MAC in young adults’) was found to enhance the occurrence of high-frequency activity at ≥80 Hz, in a dose-dependent manner and preferentially at the epileptogenic, medial temporal sites (Orihara et al., 2020). Their results infer that 2.5% and 3.0% sevoflurane (equivalent to 1.2 and 1.7 MAC in young adults) optimized classification (Orihara et al., 2020). Note that the reported conversion between percent sevoflurane to MAC was done based on the established, age-related MAC charts (Nickalls and Mapleson, 2003).

The collective evidence mentioned above encourages prospective studies of large cohorts to determine whether MI under sevoflurane anesthesia can predict seizure outcomes following focal, resective surgery and ultimately reduce the need for extraoperative, invasive evaluation. In conjunction, further studies are also warranted to investigate whether sevoflurane-induced MI may optimize the placement of intracranial electrodes for patients who definitively need extraoperative ECoG recording and/or can potentially localize the epileptogenic zone in those who fail to show interictal or ictal epileptiform discharges during weekly extraoperative ECoG recording.

### 4.2. Innovation

The main innovation of this study is the objective and rapid measurement of MI during intraoperative ECoG recording. Speaking to the latter, visual counting of interictal spikes or high-frequency oscillations is time-consuming and suffers from suboptimal inter-rater agreement. Our previous study reported that the computational time of the MI quantification for a five-minute ECoG recording was 1.2 seconds/channel, whereas that required to quantify the occurrence rate of high-frequency activity ranged from 3.7 to 89.3 seconds/channel (Kuroda et al., 2021). Thus, one may hypothesize that MI-based evaluation may provide investigators with a longer planning time than that based on the occurrence rate of high-frequency activity.

### 4.3. Mechanisms underlying sevoflurane-induced augmentation of modulation index (MI)

The MI augmentation preferentially at epileptogenic sites may be partly attributed to an increase in the magnitude and occurrence rate of interictal spike-and-wave discharges (Kuroda et al., 2021; Motoi et al., 2018). By definition, MI reflects phase-amplitude coupling between high-frequency amplitude at ≥150 Hz and the phase of a slow wave at 3-4 Hz, and it is a proxy measure for spike discharges generally containing broadband high-frequency activity immediately followed by a slow wave (Fürbass et al., 2020; Jacobs et al., 2011). Thus, sites showing *<u>large</u>* and *<u>repetitive</u>* spike discharges followed by morphologically similar slow waves at 3-4 Hz would have higher MI values than those showing *<u>small</u>* and *<u>sporadic</u>* spike discharges not necessarily accompanied by such slow waves.

While this speculation is well founded, further studies are required to definitively determine the cellular mechanisms underlying the sevoflurane-induced increase in MI or spike-and-wave discharges at epileptogenic sites. Previous data indicated that inhibition of excitatory receptors (e.g., N-methyl-d-aspartate [NMDA] receptors) and potentiation of inhibitory receptors (e.g., gamma-aminobutyric acid [GABA] receptors) play causative roles in sevoflurane-based general anesthesia (Brosnan and Thiesen, 2012; Nishikawa and Harrison, 2003). In contrast to healthy sites, epileptogenic brain regions display upregulated Na-K-Cl co-transporter-1 (NKCC1) expression, which directly increases the intracellular concentration of inhibitory Cl^-^ ions (Aronica et al., 2007; Okabe et al., 2002; Palma et al., 2006). Consequently in this scenario, one could surmise that sevoflurane-induced GABA receptor activation paradoxically leads to Cl^-^ efflux and subsequent depolarization in neurons (Doi et al., 2021). Indeed, a previous study demonstrated that the epileptogenic lesion was characterized by a GABA-mediated, depolarized reversal potential (Conti et al., 2011).

Although MI was typically enhanced under sevoflurane, specifically at epileptogenic sites, the 2.0 MAC stage presumably elevated MI in non-seizure generating tissue, as well. Although the exact mechanism responsible for this observation remains to be determined, several explanations may account for this effect. It is plausible to expect that interictal spike-and-wave discharges may have propagated extensively from the epileptogenic to non-epileptogenic sites during such an advanced anesthetic stage. Previous studies of patients with focal epilepsy reported that interictal spike-and-wave discharges were often deactivated during wakefulness and rapid-eye-movement sleep but activated and propagated to extensive non-epileptogenic brain regions during slow-wave sleep (Malow et al., 1997; Luat et al., 2005; Bagshaw et al., 2009; Okanari et al., 2015). While spike-generating sites reportedly tend to show a larger amplitude of spike discharges than those receiving such propagations, that is not always the case and may contribute to our observations (Asano et al., 2003; Hufnagel et al., 2000). It is also possible that the ‘*2*.*0 MAC maintenance*’ stage may have augmented non-specific, sharp transients diffusely coupled with prolonged low-amplitude slow waves, mimicking a burst-suppression pattern.

### 4.4. Methodological considerations

In the present study, we believe that intraoperative ECoG signals during the ‘*first 1/3*’ period were affected by the mixed effects of propofol and remifentanil in addition to sevoflurane, but these effects most likely waned by the ‘*last 1/3*’ period. Some investigators previously inferred that the occurrence rate of high-frequency activity was increased as the residual concentration of propofol gradually faded in the brain (Zijlmans et al., 2012). Oppositely, remifentanil reportedly enhanced the occurrence rate and topography of spike discharges, in a dose-dependent manner, for seven out of 59 study patients with mesial temporal lobe epilepsy (Kjaer et al., 2017). In our current retrospective analysis, we could not strictly control the effects of propofol and remifentanil on ECoG signals. Thus, we cannot rule out the possibility that the suboptimal performance of the MI-based classification during the ‘*first 1/3*’ period was due to the mixed effects of propofol or remifentanil administration.

Apart from synergist effects of multiple anesthetic agents complicating the present results, we must also consider that sevoflurane concentration was not quantally increased but rather dynamically elevated to 2.0 MAC. Thus, it remains to be determined whether sevoflurane stably ranging around 1.5 MAC is more efficacious for localizing the epileptogenic zone than dynamically rising from 1.5 to 2.0 MAC. Based on the results of the present and prior studies (Orihara et al., 2020), one may consider a new anesthesia protocol in which the sevoflurane concentration gradually increases from 2.0% (equivalent to 1.0 MAC in young adults) in a stepwise manner; this approach is expected to secure a stable period of ECoG with sevoflurane maintained at 1.5 MAC and other concentrations-of-interest. Further studies are expected to determine the underlying etiology accounting for such inter-individual variability in the sevoflurane-induced activation of epileptiform discharges, as shown in **Figure 1**. Likewise, one needs to take into account the effect of hypocapnia on MI measurement in future studies. Hypocapnia induced by hyperventilation is generally associated with increased rates of epileptiform discharges and ictal events in patients with focal epilepsy (Guaranha et al., 2005).

Thinking about our statistical analysis, we analyzed a total of 621 intracranial electrode sites sampled from eight patients, and employing mixed model-based analyses provided us with excellent statistical power, despite the small number of study patients. Since the mixed model analysis considers each patient as a random factor, it effectively removes potential bias due to inter-individual variability, when quantifying the effect of a given anesthetic stage on MI distribution. Unfortunately, the present work did not benefit from ECoG sampling at the whole-brain level, so we could not consider the normative anatomical variability in MI. Our previous study of extraoperative ECoG recording demonstrated that the non-epileptic/healthy occipital cortex had higher MI than the other healthy lobes (Kuroda et al., 2021). In a similar fashion, we plan to build a whole-brain level, normative atlas of phase-amplitude coupling under general anesthesia, aiming to improve the diagnostic accuracy of anesthesia-based localization of the epileptogenic zone.

## Data Availability

All data produced in the present study are available upon reasonable request to the authors

## Abbreviations

ECoG: electrocorticography
EEG: electroencephalography
MI: modulation index
GABA: gamma-aminobutyric acid
MRI: magnetic resonance imaging
MAC: minimum alveolar concentration
ILAE: International League Against Epilepsy

## Acknowledgments

This work was supported by KAKENHI Grant JP19K09494 (to M.I.) and NIH grant NS064033 (to E.A.).

